# Spatial–Temporal Trajectories of County-Level Low Birthweight Rates and Contextual Determinants in the United States

**DOI:** 10.1101/2024.01.15.24301330

**Authors:** Pallavi Dwivedi, Thu T. Nguyen, Pankaj Dipankar, Carson J Peters, Xin He, Quynh C. Nguyen

## Abstract

**Background:** Infants born with low birthweight (LBW; <2,500 grams) are at increased risk of infant mortality, long-term neurologic impairment, and chronic conditions such as cardiovascular disease and diabetes. Although maternal risk factors for LBW are well documented, national and state-level estimates may mask substantial county-level heterogeneity in LBW risk and its underlying social and structural determinants. Less is known about how LBW rates vary geographically across metropolitan and non-metropolitan counties in the United States (U.S.) and how county-level contextual factors contribute to these patterns over time.

**Methods:** This longitudinal study included 18,193,145 singleton births across 3,108 U.S. counties during the study period (2016-2021). Birthweight and maternal characteristics were obtained from restricted-use National Center for Health Statistics natality files, and county-level contextual variables were derived from County Health Rankings data. Counties were classified as large metropolitan, small metropolitan, or non-metropolitan using the National Center for Health Statistics urban–rural classification scheme. Bayesian hierarchical spatial models were used to estimate smoothed standardized incidence ratios of LBW. Generalized estimating equations with negative binomial regression were applied to examine associations between county-level contextual characteristics and LBW rates over time, and interaction terms were used to assess effect modification by metropolitan status.

**Results:** LBW rates increased significantly across U.S. counties over the study period, with substantial geographic heterogeneity. After adjustment for maternal and county-level covariates, metropolitan status was not significantly associated with LBW rates, and no statistically significant interactions were observed between metropolitan status and time or other maternal or socioeconomic characteristics. Higher county-level percentages of Black women, underweight women, women who smoked during pregnancy, women receiving no prenatal care, and uninsured populations were significantly associated with increased LBW rates.

**Conclusions:** We observed substantial increase in LBW rates from 2016-2021 across the U.S. counties, with marked spatial heterogeneity that was not explained by metropolitan status. County-level socioeconomic and maternal health characteristics were key predictors of elevated LBW risk. Findings underscore the need for targeted, place-based interventions addressing structural and socioeconomic vulnerabilities, particularly in communities with high proportions of racially minoritized and uninsured populations, regardless of metropolitan status.

## BACKGROUND

In comparison with normal birthweight infants, neonates with low birthweight (< 2500 grams at birth) have a higher risk of mortality and chronic diseases such as cardiovascular disease and diabetes.^1,2,3,4^ The determinants of low birthweight include multiple gestation or carrying more than one fetus, obstetric complications, infections, smoking, drinking alcohol, drug abuse, stress, undernutrition, and chronic maternal conditions such as hypertension.^2,4,5,6^ Prenatal exposure to air pollution has also been shown to be associated with low birthweight.^2^ Previous studies have also suggested that racial discrimination is associated with increased risk of preterm and low birthweight.^7,8^ The low birthweight rate for Black people has been more than twice that for White people from 2006-2016.^9^ Though previous research has documented the effect of aforementioned contextual factors on low birthweight rates, it is not clear whether the effect of these factors on low birthweight rates varies based on the metropolitan status of counties.

The National Center for Health Statistics estimated the prevalence of low birthweight in the U.S. to be 8.3% in 2018.^10^ The Healthy People 2020 goal was to reduce the low birthweight rate to 7.8%.^11^ Previous research has suggested that pregnant women living in rural areas have higher risk of mortality and are less likely to seek or receive prenatal and postnatal care and screening as compared to women living in urban areas.^12, 13^ These rural–urban disparities may be attributable to limited health care access and reduced utilization of prenatal and postpartum health services in socioeconomically disadvantaged rural areas.

Although prior studies have documented individual-level and state-level determinants of low birthweight, fewer studies have examined how low birthweight risk evolves over time at the county level while accounting for spatial dependence and contextual factors. Moreover, it remains unclear whether rural–urban differences persist after adjusting for county-level maternal composition, socioeconomic conditions, and healthcare access. Addressing these gaps is critical for identifying high-risk areas and informing geographically targeted public health interventions.

In this study, we explored the effect of contextual factors on county-level low birthweight rates and whether the effects of socioeconomic disadvantage and maternal factors, such as marital status, live birth order, cigarette smoking, education, and race, on low birthweight rates vary by county metropolitan status. We also examined whether there was any change in low birthweight rates by metropolitan status of counties from 2016-2021 and the distribution of low birthweight risk across the counties. We hypothesized that county-level low birthweight rates would exhibit significant spatial and temporal heterogeneity, that socioeconomic disadvantage and maternal health characteristics would be associated with higher rates of low birthweight, and that these associations might differ by metropolitan status. Examination of geographical variation in low birthweight rates and associated contextual factors can help inform interventions and policies to reduce low birthweight.

## METHODS

### County-level health data

This longitudinal study utilized data on 18,193,145 singleton births across 3,108 counties for the time period of 2016-2021 in the United States (U.S.). Multiple-plurality births and singleton births with congenital anomalies were excluded from the analyses. Data on birthweight and maternal characteristics (e.g., live birth order, race, age, marital status, prenatal care, body mass index, cigarette smoking status during the first, second, or third trimester of pregnancy, maternal education) were obtained from the restricted natality files with geographical identifiers from the National Center for Health Statistics (NCHS).^14^ Data on county-level time-varying contextual variables, including the percentage of the unemployed population, the percentage of the uninsured population, the violent crime rate per 100,000 population, and the primary care provider rate per 100,000 population, were obtained from the County Health Rankings data.^13^

### Measures

Counts of low-birth-weight cases were aggregated by the county of the mother’s residence in two-year study periods (2016-2017, 2018-2019, and 2020-2021) to enable stabilization of low birthweight rates. Since several women do not give birth in their residence county, the counts of low birthweight cases were generated based on the mother’s residence county rather than the county where the birth occurred. We compared the proportion of low birthweight cases across the study periods and, within each study period, the proportions by maternal characteristics such as race, prenatal care, smoking during pregnancy, underweight, age group, body weight based on Body Mass Index (BMI), education, marital status, and live birth order using the chi-square test. To examine the effect of these maternal characteristics at the county scale on the change in low birthweight rates over time (2016-2021), data on county-level time-varying contextual variables, such as percentage of unemployed population, percentage of uninsured population, violent crime rate per 100,000 population, and primary care provider rate per 100,000 population, were generated for the two-year study periods. We also generated county-level percentages of maternal characteristics such as race, being underweight based on BMI, cigarette smoking during pregnancy, live birth order of more than one, maternal education, lack of prenatal visits, and women within the age range of 35-54 years using the NCHS data for the two-year study periods. The NCHS Urban–Rural Classification Scheme for counties was used to classify counties.^15^ The NCHS scheme is a six-category classification scheme based on county population. The six categories were combined into three: large metropolitan counties (metro areas with a population of 250,000 or more), small metropolitan counties (metro areas with fewer than 250,000 population), and non-metropolitan counties.

### Statistical Analysis

The geographic distribution of low birthweight rates for each two-year study period was examined by calculating the Standardized Incidence Ratios (SIRs). The SIRs can be calculated as the ratio of the observed to the expected number of low birthweight cases for each county. State-level low birthweight rates were used to calculate county-level expected counts of low birthweight cases by multiplying them by the county’s total number of births. County-level observed (raw) rates of low birthweight can be misleading, particularly when data are sparse due to small population size. Therefore, a Bayesian hierarchical model was used to compute smoothed SIRs, and posterior marginals of SIRs were used to generate the exceedance probability that the SIR exceeds 1 for each county during each study period.^16, 17^ The Bayesian model included a spatial random effect term, which captures the influence of neighboring counties, and a non-spatial, or unstructured, variability term. The spatial random effect term can be defined as a conditional autoregressive Gaussian process with mean equal to the mean of the spatial effects of the neighboring geographic units and precision (τ_v_). The non-spatial random effect term was normally distributed with a mean equal to zero and precision (τ_u_). Gamma (0.5, 0.0005) priors were assigned to the precision terms in the model. The R-INLA package was used to conduct Bayesian inference via the Integrated Nested Laplace Approximation (INLA) method.^18–20^ Spatial smoothing was employed to reduce instability in county-level estimates arising from small population sizes and sparse data, a common limitation in analyses of rare outcomes at fine geographic scales.

A marginal log-linear regression using generalized estimating equations (GEE) model was employed to examine the association between county-level contextual variables and low birthweight rates from 2016-2021. Examination of the distribution of low birthweight rates within each county classification (non-metropolitan, small metropolitan, and large metropolitan) revealed that the variance exceeded the mean, indicating overdispersion. Accordingly, a negative binomial regression model was specified to estimate the count of low-birthweight cases. The logarithm of the population at risk (total number of births per county) was included as an offset, allowing regression coefficients to be interpreted as rate ratios.

Beyond estimating overall associations, we sought to assess whether the relationship between county-level contextual and maternal characteristics and low birthweight rates differed by metropolitan status. Metropolitan and non-metropolitan counties differ substantially in environmental characteristics, public health infrastructure, access to health care resources, policies, and other structural constraints, which may modify the impact of socioeconomic disadvantage and maternal risk factors on birth outcomes. Therefore, interaction terms between county metropolitan status and key socioeconomic indicators, specifically county-level percentages of uninsured and unemployed populations, were included to test the hypothesis that socioeconomic disadvantage exerts differential effects on low birthweight rates across urban–rural contexts. In addition, we examined interactions between metropolitan status and time to assess whether temporal trends in low birthweight rates varied by county type. This analysis accounts for potentially differential exposure to changing economic conditions, the implementation of state and local health policies, demographic shifts, or public health shocks over the study period. We also analyzed interactions between metropolitan status, maternal race, and marital status. The rationale was to investigate whether the well-established disparities in low birthweight are more or less pronounced in metropolitan versus non-metropolitan counties, as the social, economic, and healthcare mechanisms that produce these inequities vary by geographic setting. We also tested for the mediating effect of socioeconomic status on the association between racial groups and low birthweight rates.

County-level maternal characteristics such as percentage of live birth order > 1, women aged 35–54 years, being underweight, being educated until high school or less, cigarette smoking during pregnancy, and no prenatal care visits were included in the model to assess them as predictors of low birthweight rates. Violent crime rate and primary care provider rate per 100,000 population were included as confounders in the regression model. Correlation among repeated observations within counties across the three study periods was accounted for using an unstructured working correlation matrix in the GEE framework. All predictor variables in the GEE model were standardized to have a mean of 0 and a standard deviation of 1. This study was approved by the University of Maryland Institutional Review Board (IRB).

## RESULTS

There were a total of 1,273,614 low birthweight cases, out of 18,193,145 singleton births, over the duration of 2016-2021. The percentage of low birthweight cases was similar across the first two study periods but increased in the third study period (477,971 [6.42% of total 7440664 births] in 2016-2017; 472,319 [6.56% of total 7200506 births] in 2018-2019; 323,324 [9.10%] of total 355,1975 births in 2020-2021; p < 0.001). During all the study periods, a higher percentage of infants born to Black mothers had low birthweight as compared to other racial groups (p < 0.001; eTables 1-3). The percentage of low birthweight cases was consistently higher among infants whose mothers did not receive or seek any prenatal care as compared to infants whose mothers received at least one prenatal visit (17.61% versus 6.10% for 2016-17; 17.47% versus 6.24% for 2018-19; and 19.85% versus 6.43% for 2020-21). Across all the study periods, the percentage of low birthweight newborns was higher among women who smoked during pregnancy as compared to women who did not smoke during pregnancy (12.15% versus 5.97% for 2016-17; 12.47% versus 5.78% for 2018-19; and 15.77% versus 5.86% for 2020-21). A higher percentage of infants born to mothers within the age group of less than 20 years and more than 40 years had low birthweight as compared to women in other age groups across all the study periods. Across all the study periods, a higher percentage of low birthweight infants were born to underweight women as compared to infants born to normal or obese women (10.89% versus 6.21% or 6.12% for 2016-17; 11.31% versus 6.33% or 6.30% for 2018-19; and 55.00% versus 6.54% or 6.36% for 2020-21).

Additionally, in comparison with married women, a higher percentage of infants born to unmarried women had low birthweight in all the study periods (5.08 % versus 8.60 % for 2016-17; 5.21 % versus 8.92 % for 2017-18, and 5.25 % versus 9.05 % for 2020-21). A higher percentage of low birthweight infants were born to women with live birth order of one as compared to women with live birth order of more than one during the study periods of 2016-17 (7.51 % versus 5.73 %) and 2018-19 (7.66 % versus 5.86 %). There was an increase in the percentage of low birthweight infants in all types of counties in the study period of 2020-21 as compared to the study periods of 2016-17 and 2018-19 (9.34 % versus 6.61 % versus 6.69 % for non-metro counties; 9.01 % versus 6.42 % versus 6.54 % for small metro counties and 9.07 % versus 6.39 % versus 6.54 % for large metro counties). At the county scale, the median low birthweight rate was 6.17 per 100 births in 2016-2017, 6.29 per 100 births in 2018-2019, and 8.92 per 100 births in 2020-2021. Collectively, these patterns indicate persistent and widening disparities in low birthweight associated with maternal race, prenatal care utilization, smoking during pregnancy, and nutritional status across all study periods.

### Geographic distribution of low-birth-weight rates

Figure 1 presents the smoothed Standardized Incidence Ratios (SIRs) for low birthweight and the Bayesian exceedance probability for SIRs greater than 1 for the three study periods. SIRs greater than 1 indicate a higher than expected or state-level risk of low birthweight, and SIRs less than 1 represent a lower than expected risk of low birthweight. We observed an elevated risk of low birthweight, with an exceedance probability of more than 0.95, in several counties in California, Oregon, Washington, Nevada, Arizona, Utah, Wyoming, Idaho, Colorado, New Mexico, Minnesota, and in the southern, midwestern, and northeastern states over the three study periods (Figure 1). These spatial patterns were largely consistent across study periods, suggesting persistent geographic clustering of elevated low birthweight risk rather than transient fluctuations.

**Figure 1:**
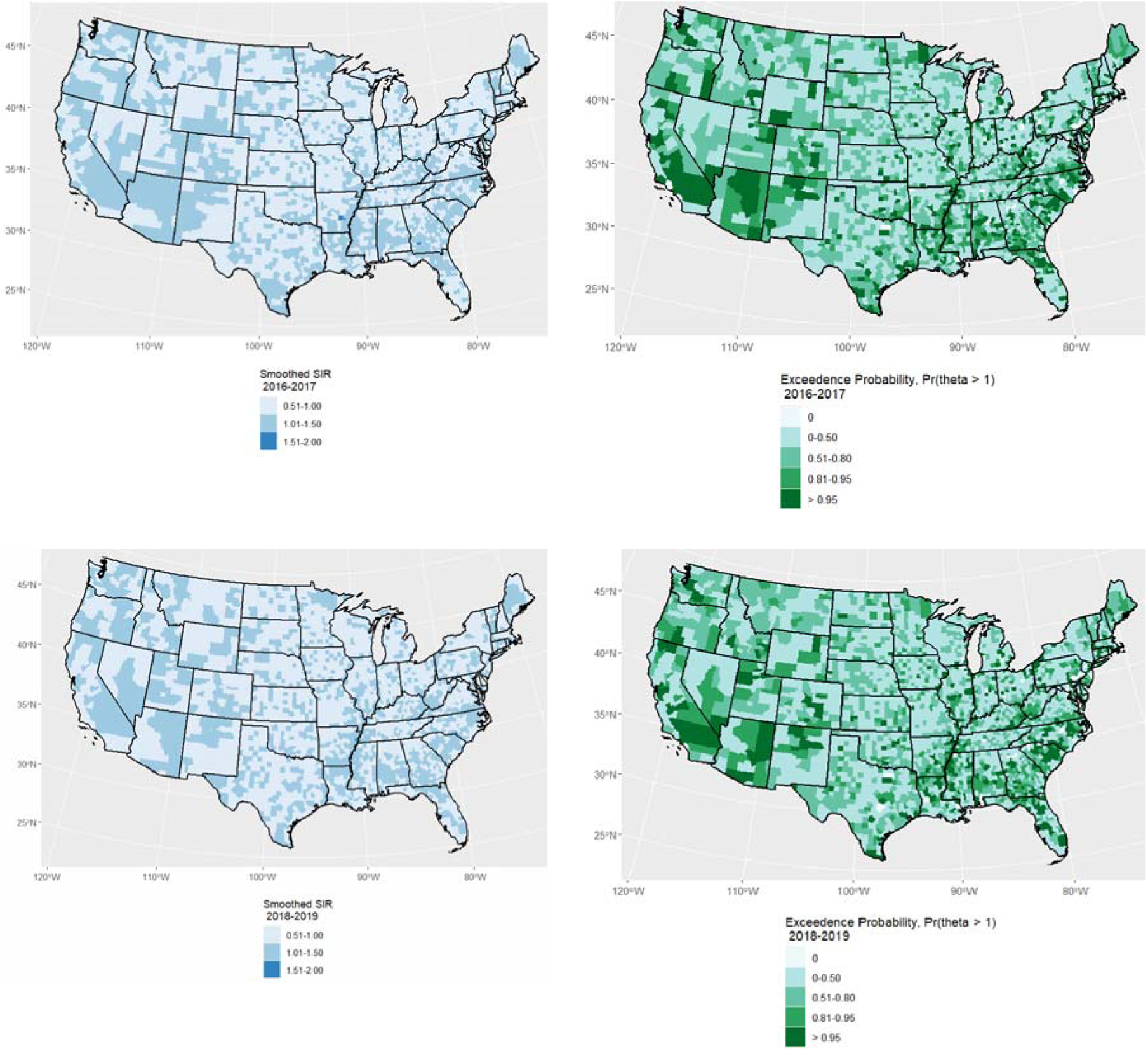
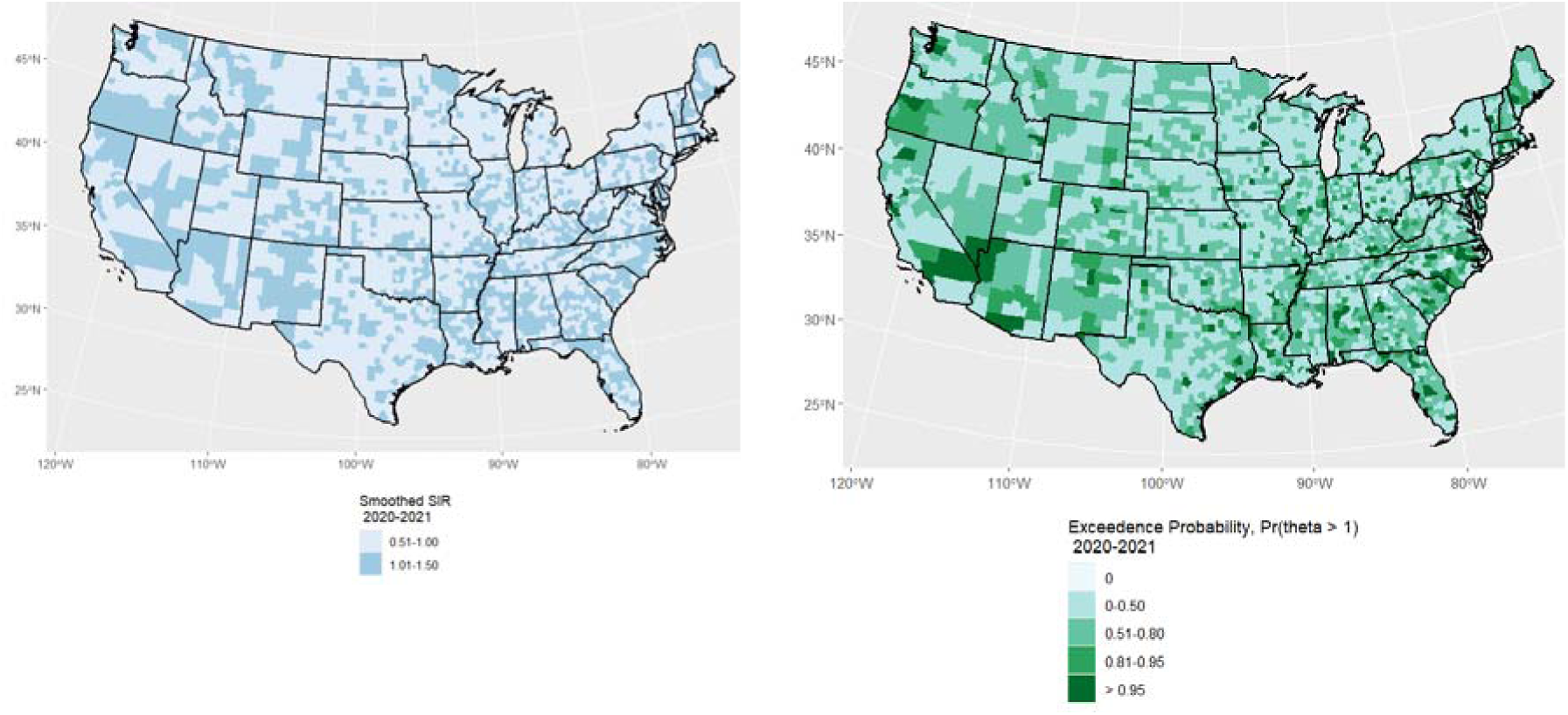
Smoothed Standardized Incidence Ratios (SIRs) of low birthweight and exceedance probability of low-birth-weight SIR > 1 for counties across the U.S. for 3 study periods (2016-2017, 2018-2019, and 2020-2021) County-level maps display the spatial distribution of smoothed SIRs, where values greater than 1 indicate higher-than-expected incidence of low birthweight relative to the state average. Accompanying exceedance probability maps show the posterior probability that the SIR exceeds 1, with higher probabilities indicating stronger evidence of elevated risk. Results are presented separately for each study period to illustrate temporal changes in the geographic pattern of low birthweight risk.

### Contextual variables associated with low birthweight rates

Table 1 presents the incidence rate ratios (IRR) for low birthweight for each predictor variable after adjusting for all the other covariates. After adjusting for all the covariates, one standard deviation increase in the county-level percentage of Black women (incidence rate ratio, IRR = 1.14, 95% CI = 1.13, 1.15) was associated with higher low birthweight rates. One standard deviation increase in the percentage of women who did not obtain any prenatal care, women who were underweight based on their BMI, and women who smoked cigarettes during pregnancy was associated with increased low birthweight rates (IRR for no prenatal care = 1.03, 95% CI = 1.02, 1.03; IRR for underweight based on BMI = 1.11, 95% CI = 1.10, 1.12; IRR for smoking during pregnancy = 1.05, 95% CI = 1.04, 1.06, respectively). One standard deviation increase in the county-level percentage of uninsured women was associated with a significant increase in the low birthweight rate (IRR = 1.04, 95% CI = 1.03, 1.05). Excluding the county-scale percentages of the uninsured and unemployed populations did not weaken the effect of the percentage of the Black population on low birthweight rates, nor did it change the effect of the percentage of other maternal racial groups on low birthweight rates.

**Table 1:**
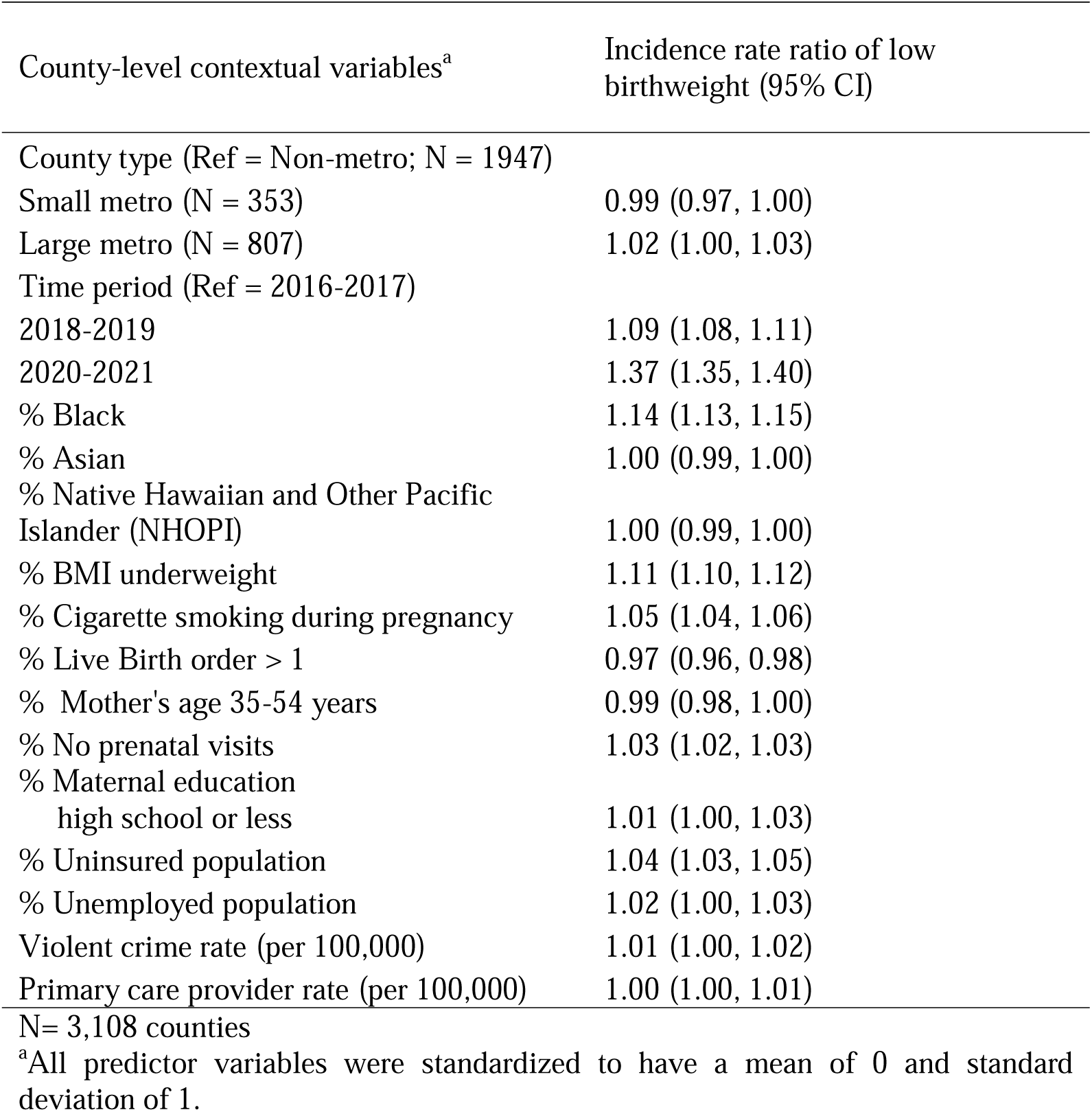
Association of county-level contextual variables with low-birth-weight rates during the study period (2016-2021).

| County-level contextual variables<sup>a</sup> | Incidence rate ratio of low birthweight (95% CI) |
| --- | --- |
| County type (Ref = Non-metro; N = 1947) |  |
| Small metro (N = 353) | 0.99 (0.97, 1.00) |
| Large metro (N = 807) | 1.02 (1.00, 1.03) |
| Time period (Ref = 2016-2017) |  |
| 2018-2019 | 1.09 (1.08, 1.11) |
| 2020-2021 | 1.37 (1.35, 1.40) |
| % Black | 1.14 (1.13, 1.15) |
| % Asian | 1.00 (0.99, 1.00) |
| % Native Hawaiian and Other Pacific Islander (NHOPI) | 1.00 (0.99, 1.00) |
| % BMI underweight | 1.11 (1.10, 1.12) |
| % Cigarette smoking during pregnancy | 1.05 (1.04, 1.06) |
| % Live Birth order > 1 | 0.97 (0.96, 0.98) |
| % Mother's age 35-54 years | 0.99 (0.98, 1.00) |
| % No prenatal visits | 1.03 (1.02, 1.03) |
| % Maternal education high school or less | 1.01 (1.00, 1.03) |
| % Uninsured population | 1.04 (1.03, 1.05) |
| % Unemployed population | 1.02 (1.00, 1.03) |
| Violent crime rate (per 100,000) | 1.01 (1.00, 1.02) |
| Primary care provider rate (per 100,000) | 1.00 (1.00, 1.01) |

N= 3,108 counties
<sup>a</sup>All predictor variables were standardized to have a mean of 0 and standard deviation of 1.

Compared to the baseline study period of 2016-2017, there was a significant increase in low birthweight rates in the study periods of 2018-2019 and 2020-2021 in all the counties after adjusting for all the covariates (2018-2019: IRR = 1.09, 95% CI = 1.08, 1.11; 2020-2021: IRR = 1.37, 95% CI = 1.35, 1.40). We did not observe a statistically significant effect of county type (large or small metropolitan counties versus non-metropolitan counties) on low birthweight rates after adjusting for all covariates. There was no statistically significant interaction between county type and any maternal or other county-scale characteristics, or between county type and time. Figure 2 presents the distribution of low birthweight rates for large metropolitan, small metropolitan, and non-metro counties over the duration of the study.

**Figure 2:**
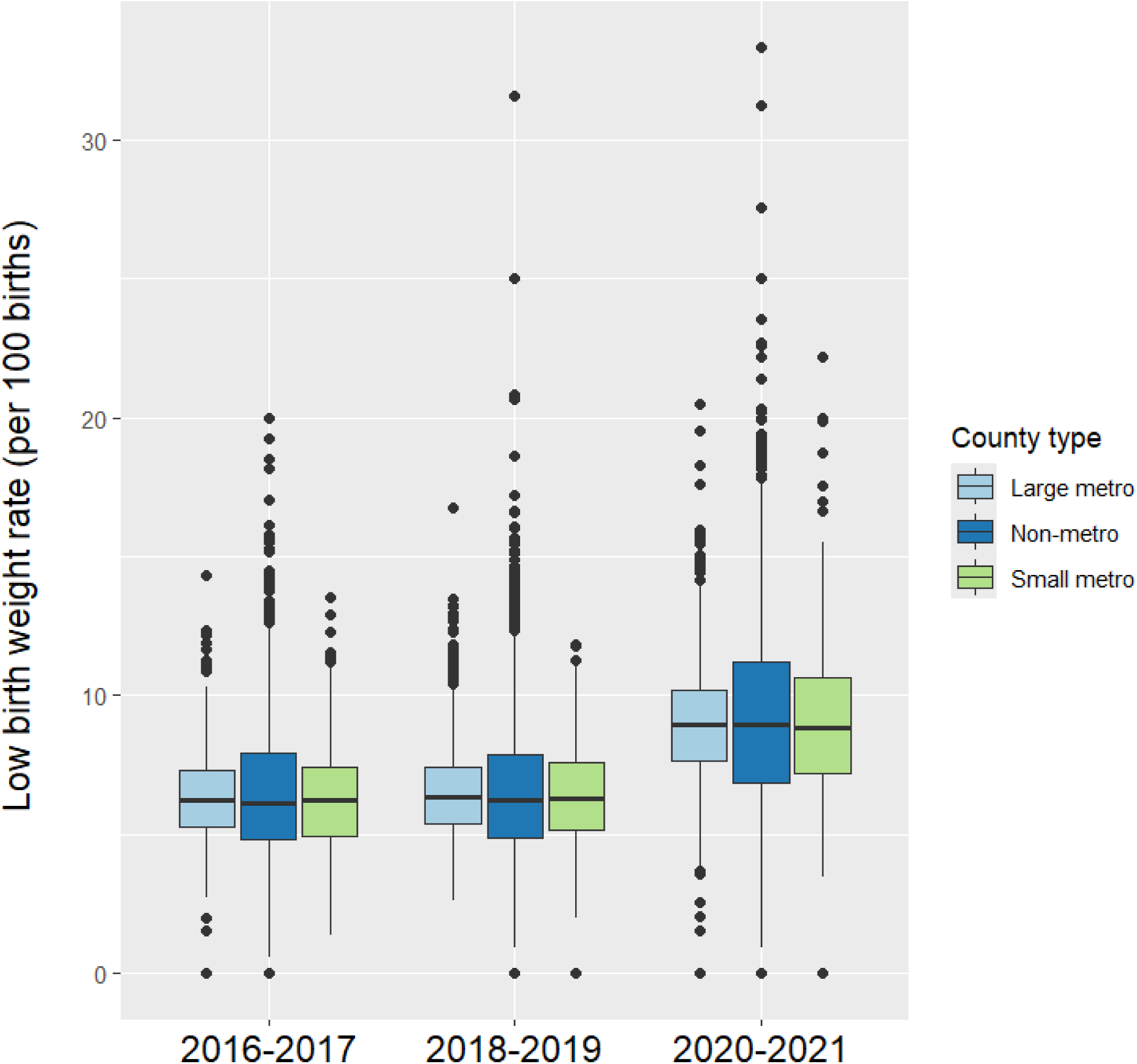
Distribution of low-birth-weight rates for non-metropolitan, small metropolitan, and large metropolitan counties over the duration of the study.

## DISCUSSION

This study examined county-level trajectories of low birthweight rates in the U.S. during the six-year period from 2016-2021 across metropolitan and non-metropolitan counties. There was a significant increase in low birthweight rates across all counties during the study period. Contrary to common assumptions about rural-urban disparities, we did not observe a statistically significant difference in low birthweight rates across counties by metropolitan status. Importantly, the absence of significant differences by metropolitan status after adjustment suggests that structural and socioeconomic conditions, rather than rural or urban location per se, may be more salient drivers of low birthweight risk.

A high county-level percentage of Black women was associated with a greater risk of low birthweight rates after adjusting for county-level demographic and socioeconomic maternal characteristics. This association may reflect the cumulative effects of structural racism, including residential segregation, differential exposure to neighborhood deprivation, and inequities in access to high-quality healthcare and social resources.^21–22^ Women who lack health insurance are less likely to obtain adequate prenatal care or schedule regular healthcare visits. In our study, we found a statistically significant positive association between the county-level percentage of the uninsured population and the lack of prenatal visits and low birthweight rates. A previous study, conducted by Taylor et al. suggested that women with Medicaid or no insurance are less likely to seek preventive care and have adverse birth outcomes such as preterm birth, preeclampsia, and low birthweight as compared to women with commercial insurance ^23^.

In addition to inadequate access to health screening or care, other factors that might affect birth outcomes include health behavior and chronic stress. Chronic stress might be another critical determinant that links structural inequities to adverse birth outcomes. Being subject to unfair judgment or treatment in the social or professional setting might increase the risk of adverse birth outcomes in women as a result of several physiological pathways linked to stress. ^24–26^ The experiences of economic instability, discrimination, and neighborhood disadvantage can activate sustained neuroendocrine and inflammatory responses, which have been linked with fetal growth restriction and low birthweight LBW rates. Furthermore, we did not find any mediating effect of socioeconomic status on the association between county scale percentage of Black women and women of other racial groups and low birthweight rates. Studies conducted by Rubin and Alhusen et al. found that maternal sociodemographic characteristics, health behavior, and prenatal care do not adequately account for disparities in birth outcomes among Black and White women^27,28^.

We observed a statistically significant positive association between a high percentage of underweight women based on their BMI and women who smoked cigarettes during the first, second, or third trimester of pregnancy and low birthweight rates. At the county level, these interconnected factors may contribute to cumulative risk environments that shape low birthweight patterns over time. Our findings support a multilevel public health framework in which structural, healthcare, and behavioral determinants interact to produce population-level disparities.

The sharp increase in low birthweight rates observed during the 2020-2021 period may reflect broader societal disruptions, including the COVID-19 pandemic. Pandemic-related stress, reduced access to prenatal care, economic instability, and changes in healthcare delivery may have disproportionately affected pregnant individuals in vulnerable communities, exacerbating existing disparities.

### Study Strengths and Limitations

A key strength of this study is large sample size and the assessment of county-level health outcomes across multiple social determinants, enabling a deeper understanding of population health disparities. This work illuminates county-level health variations that may be masked in state or national analyses and enhances the opportunity to inform targeted public health interventions. The use of Bayesian spatial modeling allowed for more reliable county-level estimates and enhanced identification of geographic areas with persistently elevated risk. We examined the effect of both individual-level and county-level maternal characteristics on low birthweight rates. A limitation is the inability to account for policy changes over time that affect health outcomes, for example, changes in access to healthcare coverage. Associations observed at the county level may not directly translate to individual-level risk, and findings should be interpreted within the context of ecological inference.

Future research should build on these findings by examining finer spatial scales and policy-relevant mechanisms that may underlie county-level heterogeneity in low birthweight risk. Analyses at the sub-county level, such as census tracts or neighborhoods, could help identify localized clusters of elevated risk that are obscured in county-level analyses and better capture residential segregation, neighborhood deprivation, and environmental exposures. Additionally, integrating measures of structural racism, chronic stress, environmental hazards, and health care quality may clarify pathways linking contextual disadvantage to adverse birth outcomes. Finally, mixed-methods or community-engaged research could complement quantitative modeling by identifying barriers to care and intervention opportunities in counties with persistently high low birthweight risk, informing more effective, place-based strategies to reduce disparities.

## CONCLUSIONS

This study demonstrates substantial and increasing county-level heterogeneity in low birthweight risk across the U.S., driven by maternal health characteristics and socioeconomic conditions rather than metropolitan status alone. Addressing low birthweight will require place-based strategies that integrate improvements in prenatal care access, insurance coverage, nutritional support, and efforts to mitigate the effects of structural racism. Such approaches are essential for reducing persistent geographic and racial disparities in birth outcomes.

## DECLARATIONS

### Ethics approval and consent to participate

This study utilized data on birth weight and maternal factors from the restricted natality files, which were obtained from the National Center for Health Statistics (NCHS) upon request. The data contains geographical location for each birth but no individual identifying information. This study was approved by the University of Maryland Institutional Review Board (IRB).

### Consent for publication

The datasets utilized in this study do not contain any individual identifying information. Administrative permission was not required to publish the findings from the data.

### Availability of data and materials

The datasets utilized in this study with geographical identifiers were obtained from the National Center for Health Statistics (NCHS) upon request.

### Author contributions

Pallavi Dwivedi: Conceptualization of research queries, statistical analyses, and wrote the initial draft of the manuscript. Thu Nguyen, Quynh Nguyen, Xin He: Refinement of research queries, editing, and revising the manuscript. Pankaj Dipankar, Carson J Peters: Editing and revising the manuscript.

## Data Availability

All data utilized for analyses in the manuscript can be obtained from National Center for Health Statistics. Restricted-Use Vital Statistics Data. (2021). https://www.cdc.gov/nchs/nvss/nvss-restricted-data.htm

## ABBREVIATIONS

LBW: low birthweight
U.S: United States
NCHS: National Center for Health Statistics
BMI: Body Mass Index
SIR: Standardized Incidence Ratio
GEE: Generalized Estimating Equations
IRB: Institutional Review Board
IRR: Incidence Rate Ratios

## Supplementary tables

**eTable 1:** Compare proportion of low birthweight cases for all the maternal characteristics for the 2016-2017 study period (N = 7440664) * p < 0.05.

| Maternal characteristics<br>(Individual birth as the<br>unit of analysis) | low birthweight<br>< 2500 g<br>N = 477971<br>Count (%) | Normal birthweight<br>≥ 2500 g<br>N = 6962693<br>Count (%) | p-value obtained from<br>Chi square test |
| --- | --- | --- | --- |
| *Racial group* |  |  |  |
| White (N = 5472449) | 293726 (5.37 %) | 5178723 (94.63 %) | < 0.001* |
| Black (N = 1192558) | 131580 (11.03 %) | 1060978 (88.97 %) |  |
| Asian (N = 516300) | 34757 (6.73 %) | 481543 (93.27 %) |  |
| American<br>Indian/Alaskan native<br>(N = 66977) | 4468 (6.67 %) | 62509 (93.33 %) |  |
| Native Hawaiian and<br>Other Pacific Islander<br>(N = 18358) | 1173 (6.39 %) | 17185 (93.61 %) |  |
| Multiple race (N =<br>174022) | 12267 (7.05 %) | 161755 (92.95 %) |  |
| *Prenatal visits (N excluding missing data for prenatal visits = 7246646)* |  |  |  |
| No prenatal visits<br>(N = 121741) | 21440 (17.61 %) | 100301 (82.39 %) | < 0.001* |
| One or more prenatal<br>visits<br>(N = 7124905) | 434507 (6.10 %) | 6690398 (93.90 %) |  |
| *Smoking status during pregnancy (N excluding missing data for smoking status = 7405779)* |  |  |  |
| Smoking (N = 523390) | 63570 (12.15 %) | 459820 (87.85 %) | < 0.001* |
| No smoking (N =<br>6882389) | 410706 (5.97 %) | 6471683 (94.03 %) |  |
| *Age group* |  |  |  |
| < 20 years (N = 396812) | 34588 (8.72 %) | 362224 (91.28 %) | < 0.001* |
| 20-24 years (N = 1512076) | 107635 (7.12 %) | 1404441 (92.88 %) |  |
| 25-29 years (N = 2175221) | 129804 (5.97 %) | 2045417 (94.03 %) |  |
| 30-34 years (N = 2092320) | 118381 (5.66 %) | 1973939 (94.34 %) |  |
| 35-39 years (N = 1036603) | 68109 (6.57 %) | 968494 (93.43 %) |  |
| 40-44 years (N = 212528) | 17829 (8.39 %) | 194699 (91.61 %) |  |
| 45-54 years (N = 15104) | 1625 (10.76 %) | 13479 (89.24 %) |  |
| *Body weight based on BMI (N excluding missing data for body weight based on BMI = 7247271)* |  |  |  |
| Underweight (N = 251732) | 27422 (10.89 %) | 224310 (89.11 %) | < 0.001* |
| Normal body weight (N = 3184403) | 197692 (6.21 %) | 2986711 (93.79 %) |  |
| Overweight/Obese (N = 3811136) | 233323 (6.12 %) | 3577813 (93.88 %) |  |
| *Education (N excluding missing data for education = 7346434)* |  |  |  |
| High school or less (N = 2878731) | 225338 (7.83 %) | 2653393 (92.17 %) | < 0.001* |
| More than high school (N = 4467703) | 245008 (5.48 %) | 4222695 (94.52 %) |  |
| *Marital status (N excluding missing data for marital status = 6984693)* |  |  |  |
| Married (N = 4179789) | 212418 (5.08 %) | 3967371 (94.92 %) | < 0.001* |
| Unmarried (N = 2804904) | 241160 (8.60 %) | 2563744 (91.40 %) |  |
| *Live birth order (N excluding missing data for live birth order = 7417868)* |  |  |  |
| 1 (N = 2858195) | 214635 (7.51 %) | 2643560 (92.49 %) | < 0.001* |
| > 1 (N = 4559673) | 261107 (5.73 %) | 4298566 (94.27 %) |  |

**eTable 2:** Compare proportion of low birthweight cases for all the maternal characteristics for the 2018-2019 study period (N = 7200506) * p < 0.05.

| Maternal characteristics<br>(Individual birth as the<br>unit of analysis) | low birthweight<br>< 2500 g<br>N = 472319<br>Count (%) | Normal birth weight<br>≥ 2500 g<br>N = 6728187<br>Count (%) | p-value obtained from<br>Chi square test |
| --- | --- | --- | --- |
| *Racial group* |  |  |  |
| White (N = 5309725) | 289046 (5.44 %) | 5020679 (94.56 %) | < 0.001* |
| Black (N = 1144722) | 130888 (11.43 %) | 1013834 (88.57 %) |  |
| Asian (N = 479098) | 33591 (7.01 %) | 445507 (92.99 %) |  |
| American<br>Indian/Alaskan native<br>(N = 66272) | 4442 (6.70 %) | 61830 (93.30 %) |  |
| NHOPI (N = 20625) | 1354 (6.56 %) | 19271 (93.44 %) |  |
| Multiple race (N =<br>180064) | 12998 (7.22 %) | 167066 (92.78 %) |  |
| *Prenatal visits (N excluding missing data for prenatal visits = 7027358)* |  |  |  |
| No prenatal visits<br>(N = 124753) | 21793 (17.47 %) | 102960 (82.53 %) | < 0.001* |
| One or more prenatal<br>visits<br>(N = 6902605) | 430577 (6.24 %) | 6472028 (93.76 %) |  |
| *Smoking status during pregnancy (N excluding missing data for smoking status = 7140415)* |  |  |  |
| Smoking (N = 446655) | 55709 (12.47 %) | 390946 (87.53 %) | < 0.001* |
| No smoking (N =<br>6693760) | 387066 (5.78 %) | 6306694 (94.22 %) |  |
| *Age group* |  |  |  |
| < 20 years (N = 345236) | 30697 (8.89 %) | 314539 (91.11 %) | < 0.001* |
| 20-24 years (N = 1379603) | 99325 (7.20 %) | 1280278 (92.80 %) |  |
| 25-29 years (N = 2085347) | 127798 (6.13 %) | 1957549 (93.87 %) |  |
| 30-34 years (N = 2075349) | 121381 (5.85 %) | 1953968 (94.15 %) |  |
| 35-39 years (N = 1075575) | 72352 (6.73 %) | 1003223 (93.27 %) |  |
| 40-44 years (N = 222947) | 18901 (8.48 %) | 204046 (91.52 %) |  |
| 45-54 years (N = 16449) | 1865 (11.34 %) | 14584 (88.66 %) |  |
| *Body weight based on BMI (N excluding missing data for body weight based on BMI = 7039412)* |  |  |  |
| Underweight (N = 221270 ) | 25022 (11.31 %) | 196248 (88.69 %) | < 0.001* |
| Normal body weight (N = 2937711) | 186029 (6.33 %) | 2751682 (93.67 %) |  |
| Overweight/Obese (N = 3880431) | 244355 (6.30 %) | 3636076 (93.70 %) |  |
| *Education (N excluding missing data for education = 7104169)* |  |  |  |
| High school or less (N = 2748131) | 219129 (7.97 %) | 2529002 (92.03 %) | < 0.001* |
| More than high school (N = 4356038) | 245062 (5.63 %) | 4110976 (94.37 %) |  |
| *Marital status (N excluding missing data for marital status = 6330013)* |  |  |  |
| Married (N = 3777430) | 196827 (5.21 %) | 3580603 (94.79 %) | < 0.001* |
| Unmarried (N = 2552583) | 227679 (8.92 %) | 2324904 (91.08 %) |  |
| *Live birth order (N excluding missing data for live birth order = 7183267)* |  |  |  |
| 1 (N = 2772623) | 212262 (7.66 %) | 2560361 (92.34 %) | < 0.001* |
| > 1 (N = 4410644) | 258624 (5.86 %) | 4152020 (94.14 %) |  |

**eTable 3:**
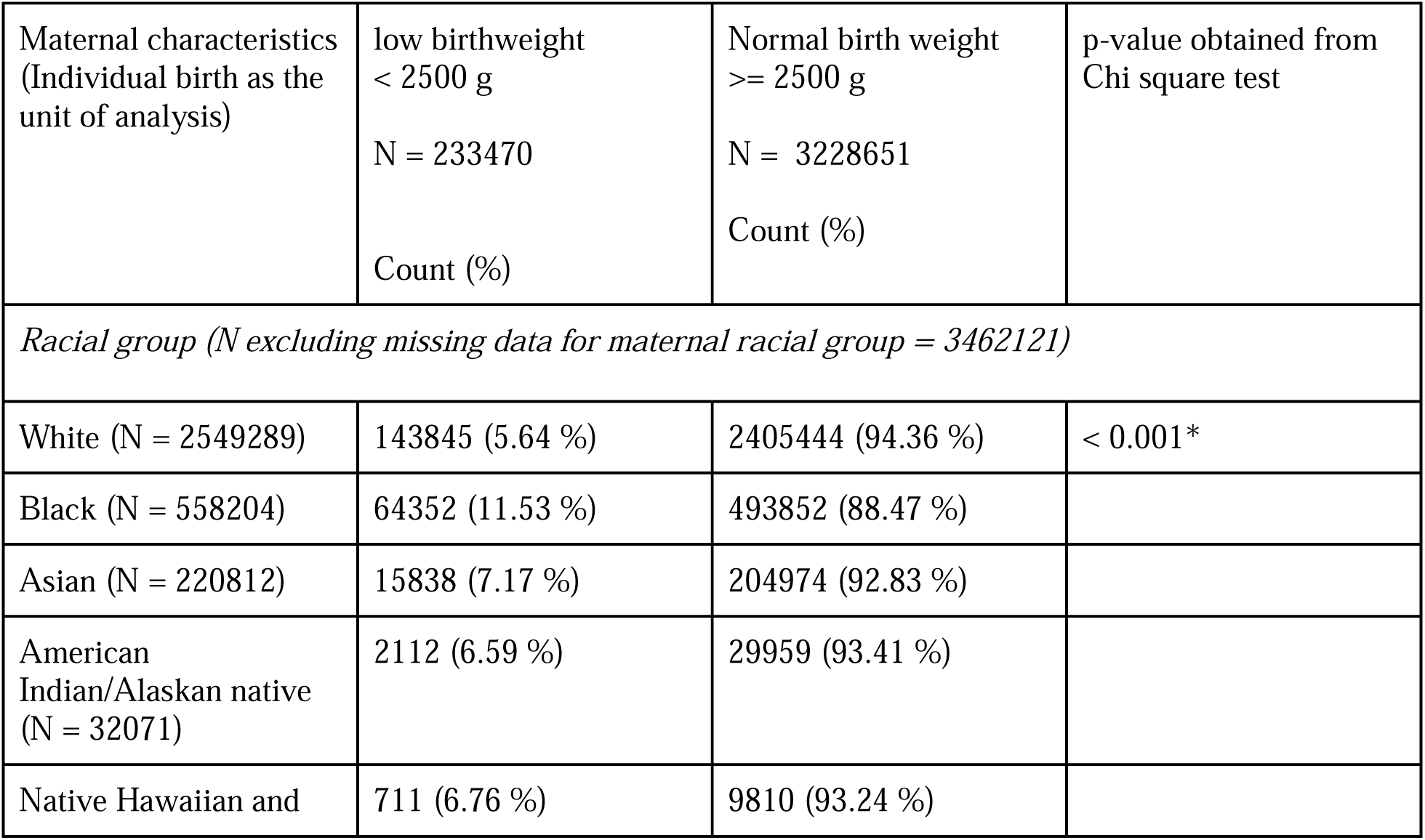
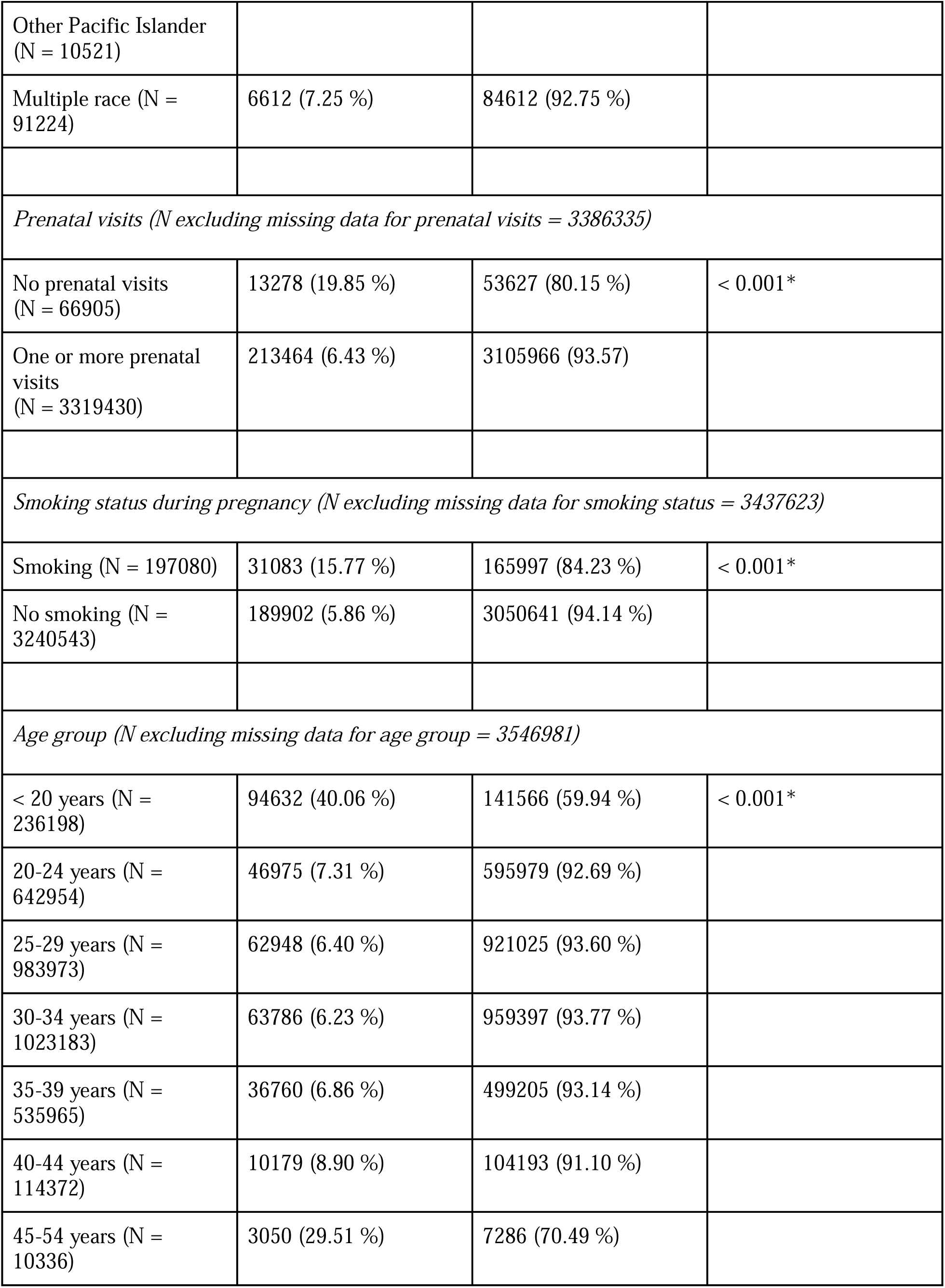
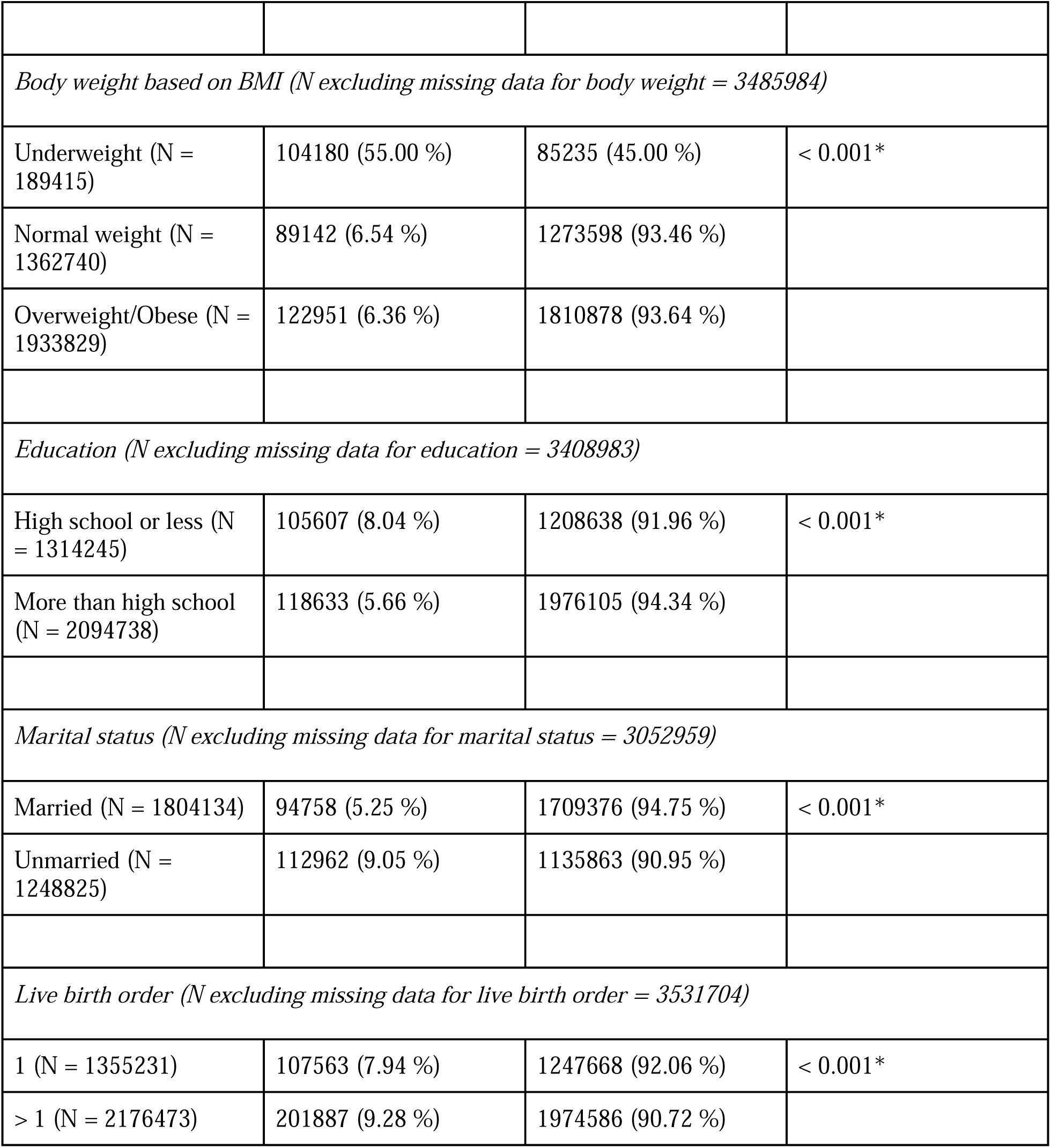
Compare proportion of low birthweight cases for all the maternal characteristics for the 2020-2021 study period (N = 3551975) * p < 0.05.

